# Role of human infection challenge studies (HICs) in drug development for respiratory syncytial virus (RSV): Systematic Review and Meta-Analysis

**DOI:** 10.1101/2024.05.24.24307680

**Authors:** Prosenjit Kundu, Mark Quinn, Elaine Thomas, Jared C. Christensen, Sima S. Toussi, Negar Niki Alami, Anindita Banerjee

## Abstract

**Background:** Human infection challenge studies (HICs) are powerful in establishing early proof-of-concept for experimental drugs and understanding disease pathogenesis. A comprehensive assessment of HICs will allow to understand the viral load dynamics and symptom score kinetics of respiratory syncytial virus (RSV) and facilitate drug development for RSV.

**Methods:** In this study, we conducted a systematic search of double-blind, placebo-controlled RSV HICs using Biosis Previews, Embase, Ovid MEDLINE, PubMed, ClinicalTrials.gov and EudraCT from 1990 to August 2023. We estimated viral load (VL) and symptom related measures in placebo and relative mean reduction (RMR) of the measures in the experimental drug compared to placebo. The primary outcomes are RMR of VL AUC and RMR of total symptom score (TSS) AUC, and the secondary outcomes are mean placebo VL area under curve (AUC), mean placebo VL at peak, time to mean placebo peak VL, mean placebo TSS AUC and time to mean placebo peak TSS. We used random-effects meta-analysis, except for time to mean peak VL and time to mean peak TSS, where descriptive statistics were summarised.

**Findings:** Number of studies varied across measures, from 4 (144 subjects in total) to 7 (247 subjects in total). Overall, relative mean reductions of 54% (95% CI: 32% – 76%, I^2^ = 91%) and 76% (95% CI: 61% – 91%, I^2^ = 21%) are observed in VL area under curve (AUC) and TSS AUC, respectively.

**Interpretation:** Assessment based on our primary outcomes showed, on average, a 54% reduction in viral load (VL) area under the curve (AUC) with significant heterogeneity between these studies. In contrast, the total symptom score (TSS) AUC showed a greater average reduction of 76%, with much lower heterogeneity indicating more consistent results across studies for this measure. Our findings inform researchers on disease course and VL kinetics, critical data needed for designing RSV treatment studies and understanding implications in clinical practice.

**Funding:** This study was supported by Pfizer Inc.

**Research in Context:** *Evidence before this study:* There are a few human challenge infection studies (HICs) that report measures related to viral load and symptom trajectory and are important for designing future clinical studies for RSV treatment. However, there is no systematic review of these HICs to elucidate the viral load kinetics and symptom dynamics and their interrelation with respect to time of achieving peak values in an integrated framework.

*Added value of this study:* We perform a systematic review of HICs and leverage information on available viral load and symptom specific outcomes into a meta-analysis framework. We provide overall estimates of the outcomes from a random-effects meta-analysis and assess the heterogeneity using the I^2^ measure. The comprehensive analysis provides insight into viral load kinetics and symptom dynamics and their temporal relationship in an adult population. We report prediction intervals of the outcomes at a study-level for a future study and reference interval, which provides a range at the individual-level for a new subject recruited in a future study. Further, we compare the total symptom score at peak and viral load at peak with Influenza and SARS-CoV-2.

*Implications of all the available evidence:* The systematic review and meta-analysis of RSV HIC studies offer a more precise estimate for viral load and symptom related measures, assess heterogeneity across studies and allude to potential sources of heterogeneity. The study provides a more nuanced understanding of amount of viral load and disease severity (measured through symptoms) over time and at peak in placebo, and how effective the experimental drugs are in viral load reduction and symptom alleviation. Further, the study helps to understand the clinical profile of RSV by comparing peak VL and peak total symptom score from other viral diseases, SARS-CoV-2 and Influenza. Finally, the findings from this study have the potential to inform modelling and sample size estimations for studies in patient populations (at risk adults and paediatrics) where such intense longitudinal viral load data are not available.

## Introduction

Respiratory Syncytial Virus (RSV) remains a major global health concern, particularly among vulnerable populations, including infants, the elderly, and individuals with underlying medical conditions^1,2^. In the United States and globally, RSV infection continues to cause severe respiratory illnesses in high-risk populations, often requiring hospitalisation and posing a significant burden on healthcare systems^3-7^. In response to this ongoing public health challenge, numerous randomised clinical trials have been conducted to assess the efficacy of various anti-viral treatments for RSV as a proof of mechanism. However, the landscape of RSV treatment is complex and remains an unmet medical need, with a wide array of interventions being tested, and outcomes that vary across studies. Data is limited in both adult and paediatric literature regarding measures related to viral load and symptom trajectory to understand the correlation of symptom trajectory and viral load (VL) for RSV. And what is available primarily focuses on the sickest population, those hospitalised with RSV^8^, and who may be late in their illness, and rarely are samples collected multiple times or with the frequency needed to characterise viral load kinetics. Thus, viral load may have already peaked by the time samples are collected and not be informative of how VL correlates with the clinical course in observational and therapeutic trials. In this study, we aim to fill the knowledge gap by providing a comprehensive analysis of viral load and symptom related measures, combining multiple challenge studies to understand viral load kinetics and symptom dynamics and thereby explicate how viral load correlates with symptom trajectory in an adult population.

Human infection challenge studies (HICs), also known as controlled human infection models (CHIM), involve intentionally exposing participants to a pathogen to test the effectiveness of treatments or vaccines^9^. For drug development, it is important to assess proof of mechanism and assess signal of efficacy early in development. A human infection challenge study allows us to understand the effect of the drug early during development and establish proof of mechanism.^10^ By running these standard human infection challenge studies, it is feasible to gauge some sign of efficacy before running larger late-phase studies and de-risk later development. For RSV, several companies have studied investigational drugs using the available human challenge model.

In this manuscript, we provide a comprehensive assessment of placebo measures and treatment efficacy reported in RSV human infection challenge studies. By conducting meta-analysis^11,12^ a widely popular statistical approach for combining studies, we seek to synthesise the available evidence, elucidate RSV VL kinetics and characterise the effectiveness of various interventions for RSV based on data from the HICs. The main objective of this meta-analysis is to summarise overall placebo measures and treatment responses for outcomes from multiple HICs published in literature or/and online clinical databases. The primary outcomes include relative mean reduction (RMR) of VL area under curve (AUC) and RMR of total symptom score (TSS) AUC, and the secondary outcomes include mean placebo VL area under curve (AUC), mean placebo VL at peak, time to mean placebo peak VL, mean placebo TSS AUC and time to mean placebo peak TSS. Random-effects meta-analysis is implemented, except for time to mean peak VL and time to mean peak TSS, where descriptive statistics are summarised. The estimated overall measures in placebo from this meta-analysis would inform the design of future RSV clinical trials. Further, as a secondary objective, we compare the placebo measures for some of the outcomes with those from published literature

for influenza and SARS-CoV-2^13,14^, to understand the clinical profile of RSV. The challenged influenza virus subtype and lineage was influenza A/Wisconsin/67/2007 (H3N2)^13^ and the challenged SARS-CoV-2 virus was Victoria/01/2020 of lineage B^14^, which is a pre-alpha virus with lineage B.1.1.7.

## Methods

### Ethics

Ethical approval was not required for this systematic review and meta-analysis, as no primary data involving human or animal subjects were collected. The data and analyses are based on previously published research.

We follow the Preferred Reporting Items for Systematic Reviews and Meta-Analyses (PRISMA) 2020 statement^15^ to report our analyses.

### Search Strategy/Study Eligibility

Following the search strategy according to the PRISMA 2020 statement^15^, the search started from clinical databases and literature websites with the key words “respiratory syncytial virus”, “RSV”, “healthy volunteers”, “healthy patients”, “healthy participants”, “healthy adults”. The search strategy was limited to English language papers only and publications from 1990 to August 2023, with any pharmaceutical product as the intervention. The websites ClinicalTrials.gov and EudraCT were searched with the key words “respiratory syncytial virus”, “RSV”, “human challenge”, “viral challenge”. We excluded 1) studies that were duplicate, 2) studies that were not human infection challenge studies, 3) studies that looked at pre-exposure prophylaxis treatment (studies where treatment, e.g., vaccine, is assigned before inoculation to challenge virus), 4) studies that used assays other than RT-qPCR and 5) studies where data is unavailable based on the outcomes related to viral load we considered in our analyses. We list them in the Statistics section. In Figure 1, we provide a PRISMA 2020 flow diagram to reflect the search strategy. In addition to outcomes, baseline demographics such as age, sex, viral load assay, and infection rate were also examined. The search strategy was independently carried out by four reviewers (PK, MQ, ET and AB).

**Figure 1:**
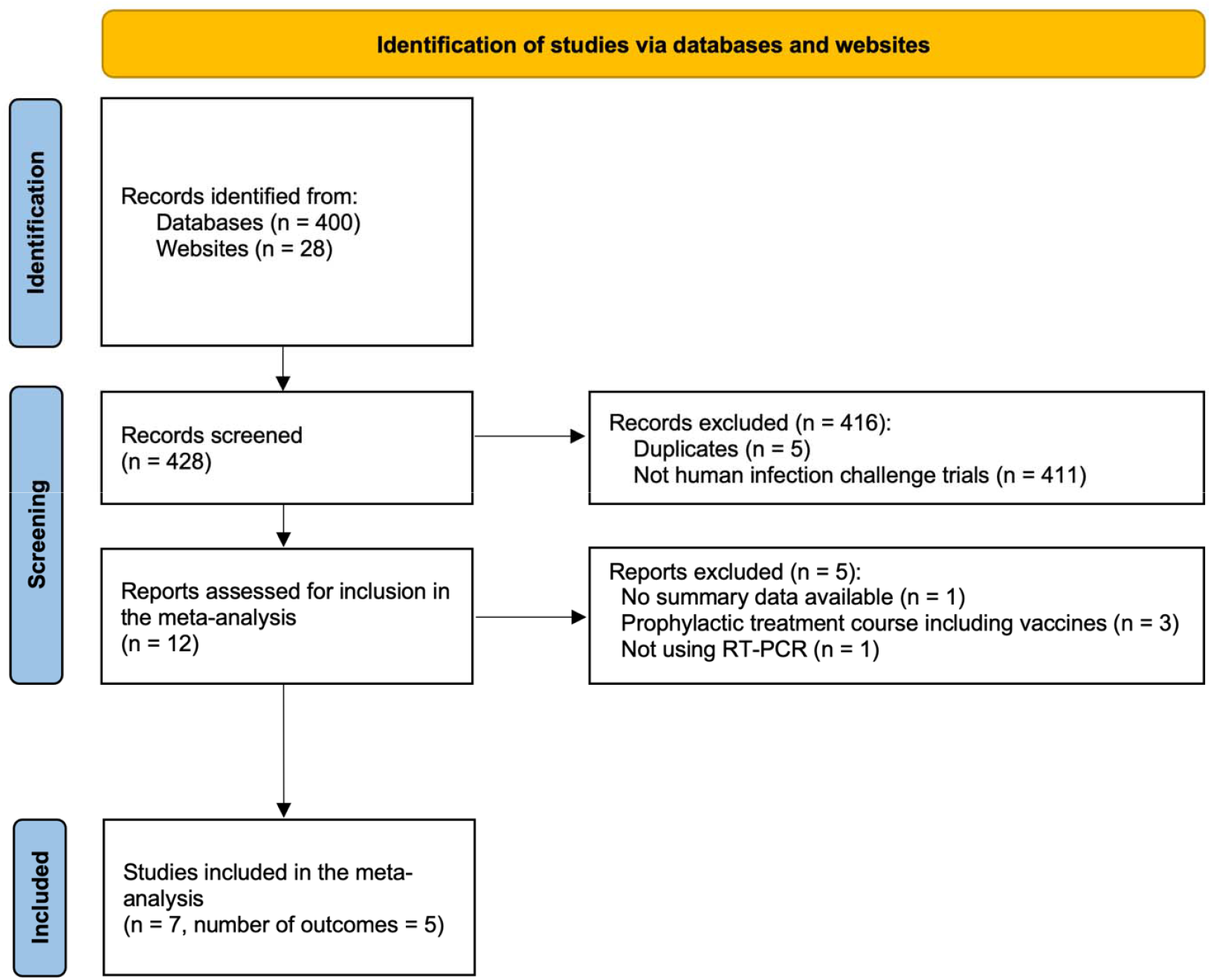
PRISMA Flow Diagram. Note: The meta-analysis is performed for 5 of the 7 study outcomes. Descriptive statistics are reported for the other 2 outcomes.

### Assessment of Risk of Bias

Bias from each of the studies is assessed independently by four reviewers based on Cochrane’s domain-based criterion. The risk of bias is evaluated using the visualisation tool robvis^16^.

### Statistics

We considered seven different outcome measures, broadly classified into viral load (VL) specific measures and symptom specific measures. VL specific measures included (1) relative mean reduction (RMR) of VL area under the curve (AUC), (2) mean placebo VL AUC where the AUC is calculated from the first dose until the end of the quarantine period (window length of ∼12-13 days for the studies included), (3) mean placebo VL at peak, (4) time (in days) to mean placebo peak VL. Symptom specific measures included (5) RMR of total symptom score (TSS) AUC, (6) mean placebo TSS AUC where the window length is similar to mean placebo VL AUC in (2), and (7) time (in days) to mean placebo peak TSS. The primary outcomes considered in our study are RMR of VL AUC and RMR of TSS AUC, and the rest are considered as secondary outcomes. The total symptom score (TSS) reported in the studies was evaluated by summing the scores from individual symptoms based on a 10-symptom diary card. The 10 symptoms included runny nose, stuffy nose, sneezing, sore throat, earache, malaise (tiredness), cough, shortness of breath, headache, and muscle/joint ache/stiffness and were graded on a scale of 0-3, where grade 0 was absence, grade 1 was just noticeable, grade 2 was bothersome but did not prevent participation in activities, and grade 3 was bothersome and interfered with activities. TSS can range from 0 to 30, with a higher score indicating a higher level of illness. In our analysis, relative mean reduction (RMR) for a measure, e.g. VL AUC is calculated as (mean VL AUC in the placebo group – mean VL AUC in the experimental drug group)/(mean VL AUC in placebo group), where mean is taken over subjects in the corresponding group. Both the time variables (4) and (7) are measured from post-inoculation. We derived the estimate of RMR, (1) and (5) from the corresponding treatment and placebo estimates reported in the studies. For deriving the time variables (4) and (7), we assumed a uniform distribution for the time (in days) from RSV inoculation to first dose assignment, where the range varied from the day of inoculation to the day of first dose assignment. This assumption is applicable for the studies which did not report data on time variables (4) and (7) from inoculation but from the assignment of the first dose. In studies that reported multiple doses for treatment regimens, we chose the cohort with the highest dose for evaluating RMR measures. Further, most (n=5) of the studies report the unit of VL AUC in log10 PFUe.hr/mL (log10 PFUe.hr/mL = log (base 10) plaque-forming unit equivalents per millilitre times hour), while some (n=2) report in log10 copies.hr/mL. Since the search strategy resulted in all the studies that were conducted by hVIVO with RSV-A Memphis 37b as the challenge strain, hVIVO confirmed the equivalence between these units. We, therefore, performed the analysis without any conversion. hVIVO is a contract research organisation specialising in infectious disease vaccines and therapeutics using human infection challenge clinical trials^17^.

We constructed a database for each of the study outcome measures. The databases included the corresponding outcome measure and associated SD for all the studies passing the inclusion/exclusion criterion (see Figure 1). The database was independently reviewed by three reviewers (PK, MQ and AB). To conduct the meta-analysis, we calculated the mean and standard deviation (SD) for each of the study outcomes. Different forms of summary statistics (from which measures such as mean and SD can be derived) were reported in the literature. Some of them were reported in numbers, and others were described in figures. For the ones reported in numbers, we took them as they are, while for the figures, we used an online digitiser tool (https://apps.automeris.io/wpd) to extract the summary statistics as numbers. Briefly, the tool allowed us to digitise a figure and then overlay a grid square using the scale on the figure axes. We could then plot digital points on the figure, and the digitiser tool returns an accurate x and y value for the point using the axes. As the uncertainty measures varied across studies, we applied different techniques to derive the estimates for SD of the four study outcomes. For the studies that reported a 95% confidence interval (95% CI), we used the two-tailed inverse of Student’s t-distribution to calculate SD to take into account the small sample size (N <30) in most of the studies. This technique calculates SD from 95% CI for a mean value t using the formula: [((95% CI)/2)/ tinv(0.05; n-1)*√n], where tinv is a function of PEIP package^18^ in R software to calculate quantiles for T-distribution. We used the delta method^19^ to estimate the SD of percentage relative reduction in mean viral load AUC in the treatment group compared to placebo. This method evaluates the variance of the percentage relative reduction as a function of matrices comprised of the means and variances of the treatment and placebo viral load means based on a normal approximation. The variances of the treatment and placebo means are evaluated from their corresponding SDs, where the SDs are computed using the tinv R function as mentioned before.

A random-effects model (using restricted maximum likelihood estimation)^20^ was implemented for each of the study outcomes (except for time to mean peak VL and time to mean peak TSS) using the R package “meta” ^21^, where the constructed database associated with the respective study outcome served as an input for the function “metamean” from the “meta” package. Forest plots were generated using the function “forest.meta”. We report the prediction interval, which offers a range at the study-level for a future study and the reference interval, which provides a range at the individual-level for a new subject. We evaluate the prediction interval of the study outcomes for a new study using the formula: 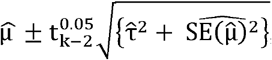, where 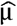 is the meta-analysis estimate, 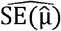 is the standard error of the meta-analysis estimate, 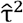 is the between study variance,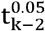 is the 97.5^th^ percentile of the t-distribution with k-2 degrees of freedom, and k denoting the number of studies^22^. Prediction interval for VL AUC, e.g., provides a range within which the mean VL AUC of a future study is expected to fall. It allows clinicians to make more informed decisions on conducting a future study and on how generalisable these results are to a target population underlying the future study. On the other hand, clinicians are often interested to know whether a patient’s outcome in a future study, e.g. VL AUC, lies within the normal range (reference interval) for healthy individuals. To that end, we evaluate the reference interval for an individual recruited in HIC using the Rshiny app: RIMeta^23^ under a random-effects model. RIMeta is an open-source user-friendly tool which takes the study-specific estimates (mean, standard error and sample size) as the input in a CSV format and reports the reference interval plots for different approaches, including frequentist, Bayesian and empirical. All three approaches assume a normal distribution of the outcome for which the reference interval is calculated^23-25^. The frequentist and the Bayesian approaches assume observations within each study are normally distributed with equal variance, while the empirical approach relaxes that assumption but instead assumes normal distribution for the population. The Bayesian approach models the common within-study variance using MCMC sampling techniques. A 95% reference interval for a particular outcome is defined to be the interval in which we would expect 95% of the individual’s outcomes from a future HIC to lie.

Most of the studies reported time to mean peak VL/TSS instead of mean time to peak VL/TSS. Mean time to peak VL/TSS is assessed by first calculating the day of peak VL/TSS for each subject and then taking the mean over days. In this case, each study would have provided a mean estimate over subjects and therefore would have fallen into the framework of traditional meta-analysis. However, on the other hand, time to mean peak VL/TSS is assessed by first calculating the mean VL/TS curve over subjects at each day and then looking at the day where the peak is achieved. Each study, therefore, provided a single observation for the time (in days) to peak VL/TSS. We treated each study as a subject and used descriptive statistics (instead of traditional meta-analysis) such as mean and median to provide overall estimates for time to mean peak VL/TSS accounting for study-level uncertainty. 95% CI for the median is calculated non-parametrically from quantiles^26^, while for the mean, we calculate it from Student’s t-distribution.

## Results

### Study Characteristics

Seven HICs^27-33^ were identified to be included in the meta-analysis (Figure 1). The search started with a total of 428 RSV studies from clinical databases and literature websites with key words stated in the Methods section. Out of the 428 studies, 11 studies were from HICs. We excluded 4 studies, based on our exclusion criterion described in the Methods section. The exclusion criterion led to 7 studies included in our analyses. These studies covered 7 different interventions in development, two of which are currently being evaluated in Phase 2 or Phase 2/3 trials, EDP-938 and Sisunatovir (RV521), respectively. All the studies are peer-reviewed, except for one^33^, whose results are published in the European Union Clinical Trial Register. The study characteristics, including demographics, are reported in Table 1. The number of subjects per study varied from 45 to 115 with a total of 544 subjects (including multiple cohorts from each study), average age per study ranged from 22.95 to 28.1 years, proportion of females varied from 24.2 to 34.2% except for one study from Johnson & Johnson^32^ which recruited male subjects only, the year of recruitment ranged from 2012 to 2018. The search criteria resulted in all studies that were conducted in the same UK centre with RSV-A Memphis-37b strain. The screening procedure was consistent across the studies, which included serosuitability, with only those individuals in the lower quartile of neutralising titres against the challenge virus strain deemed eligible. This strategy is employed to obtain an optimal infection rate^34^. Despite this, some individuals do not become infected (defined as lab confirmed RSV infection) after inoculation and this rate varied between studies (20.45 – 37.50%, Supplementary Table 1). The titre of the challenge virus in these studies is reported as 4-4.5log10 PFU/ml as determined by cell-based plaque assay, where PFU denotes plaque forming units. From Table 1, three out of the seven studies were conducted during the RSV off-season (March - October). Five of seven studies reported the distribution of baseline self-reported race categories. Participants are predominantly white (75%-91.6%) across these studies (Table 1). One of the remaining two studies, DeVincenzo et.al^29^, reported ethnicity instead of race, where 88.6% of the participants reported themselves as Caucasians, 2.3% as South Indians, and the remaining 9.1% as others (Table 1). We could not find any information about the race/ethnicity distribution from the study^33^ published in the European Union Clinical Trial Register.

**Table 1:** Study characteristics of 7 double*-*blinded, placebo-controlled human infection challenge trials^*^. ^*^ The screening process for healthy adult volunteers was consistent across studies, which included serosuitability, with only those individuals in the lower quartile of neutralising titres against the challenge virus strain deemed eligible. ^a^ Dose reported in this column indicates the highest dose among the different treatment cohorts. ^b^ N_1_ denotes the total number of subjects combining all the cohorts ^c^ N_2_ denotes the number of subjects in the combined group comprising the highest dose group and placebo ^d^ Proportions of the self-reported race/ethnic categories are calculated from the counts reported in the studies, combining the highest dose and placebo ^e^ VL denotes viral load. ^f^ RMR denotes relative reduction (in treatment compared to placebo). ^g^ AUC denotes area under curve. ^h^ TSS denotes total symptom score derived from 10-symptom diary card.

| Source | Study Period | Intervention/<br>Drug | N<sub>1</sub><sup>b</sup><br>(N<sub>2</sub><sup>c</sup>) | Mean<br>age in | Mean age<br>in years | Race/Ethnicity<sup>d</sup><br>(%) | Cohorts<br>(active | Study<br>Outcomes | VL<sup>e</sup> Assay |
| --- | --- | --- | --- | --- | --- | --- | --- | --- | --- |

**Table 1: Study characteristics of 7 double-blinded, placebo-controlled human infection challenge trials\***
|  |  | (Dose<sup>a</sup>) |  | years<br>(Range) | (Range) |  | treatment only) |  |  |
| --- | --- | --- | --- | --- | --- | --- | --- | --- | --- |
| Ahmad et al (2022)<sup>27</sup> | Oct 15, 2018 – Oct 18, 2019 | EDP-938 (5x600mg QD) | 115 (55) | 28.1 (18-55) | 34.2% | White (84.2%)<br>Black (2.6%)<br>Asian (3.9%)<br>Other (9.2%) | 2 | Mean time to peak VL<sup>e</sup>,<br>RMR<sup>f</sup> in VL<sup>e</sup><br>AUC<sup>g</sup>,<br>Peak VL<sup>e</sup> | RT-qPCR & CBI |
| Biota Pharma Europe (2018)<sup>33</sup> | Mar 23, 2016 – Dec 8, 2016 | BTA-C585 (1x600mg) | 60 (25) | 25.8 (18-64) | 33% | NA | 2 | Mean time to peak TSS<sup>h</sup>,<br>RMR<sup>f</sup> in VL<sup>e</sup><br>AUC<sup>g</sup>,<br>Peak VL<sup>e</sup> | RT-qPCR & Plaque |
| DeVince nzo et al (2014)<sup>31</sup> | Nov 2012 – June 2013 | GS-5806 (1x50mg + 4x25mg QD) | 140 (54) | 26.8 (18-45) | 24.4% | White (79.5%)<br>Black (7.7%)<br>Asian (6.4%)<br>Other (6.4%) | 4 | Mean time to peak VL<sup>e</sup>,<br>RMR<sup>f</sup> in VL<sup>e</sup><br>AUC<sup>g</sup>,<br>Peak VL<sup>e</sup> | RT-qPCR |
| DeVince nzo et al (2015)<sup>30</sup> | Mar 2014 – July 2014 | ALS-008176 (1x750mg + 9x500mg) | 62 (20) | 23.8 (18-45) | 25% | White (75%)<br>Asian (5%)<br>Other (20%) | 3 | Mean time to peak VL<sup>e</sup>,<br>RMR<sup>f</sup> in VL<sup>e</sup><br>AUC<sup>g</sup>,<br>Peak VL<sup>e</sup> | RT-qPCR |
| DeVince nzo et al (2022)<sup>28</sup> | Nov 14, 2017 – May 9, 2018 | PC786 (5x5 mg BID) | 56 (34) | 25.34 (18-55) | 31.4% | White (82.4%)<br>Asian (11.8%)<br>Other (5.9%) | 1 | Mean time to peak VL<sup>e</sup>,<br>RMR<sup>f</sup> in VL<sup>e</sup><br>AUC<sup>g</sup>,<br>Peak VL<sup>e</sup> | RT-qPCR |
| DeVince nzo et al (2020)<sup>29</sup> | July 19, 2017 – Oct 30, 2017 | RV521 (5x350mg BID) | 66 (35) | 24.6 (18-40) | 29.6% | Caucasian (88.6%)<br>South Indian (2.3%)<br>Other (9.1%) | 2 | Mean time to peak VL<sup>e</sup>,<br>Mean time to peak TSS<sup>h</sup>,<br>RMR<sup>f</sup> in VL<sup>e</sup><br>AUC<sup>g</sup>,<br>Peak VL<sup>e</sup> | RT-qPCR |
| Stevens et al (2018)<sup>32</sup> | May 7, 2015 – Oct 2, 2015 | JNJ-53718678 (7x500mg QD) | 45 (24) | 22.95 (18-45) | 0% | White (91.6%)<br>Black (4.2%)<br>Other (4.2%) | 3 | Mean time to peak VL<sup>e</sup>,<br>Mean time to peak TSS<sup>h</sup>,<br>RMR<sup>f</sup> in VL<sup>e</sup><br>AUC<sup>g</sup>,<br>Peak VL<sup>e</sup> | RT-qPCR & Plaque |

### Assessment of Risk of Bias

We reported the assessment of risk of bias for each of the HICs in Supplementary Figure 1. Biases can emerge in various areas, such as the randomisation process, outcome measurement, and other domains outlined in Supplementary Figure 1. Overall, the biases are minimal due to the studies being double-blinded, randomised controlled trials conducted at a single site by the same vendor under quarantine conditions.

### Primary outcomes

#### Relative mean reduction (RMR) of VL AUC

This analysis involved 7 studies ranging in size from 20 to 55 participants and included a total of 247 participants. Individually, the RR of mean VL AUC in the treatment group compared to placebo varied from 1% to 88% (Figure 2). The I^2^ statistic was used to assess the heterogeneity across studies and was equal to 91% (p < 0.01, Chi-squared test with 6 degrees of freedom), indicating a considerable degree of heterogeneity (Figure 2). Under a random effects model, the RMR of mean VL AUC was 54% (95% CI: 32% – 76%) compared to placebo (Figure 2).

**Figure 2:**
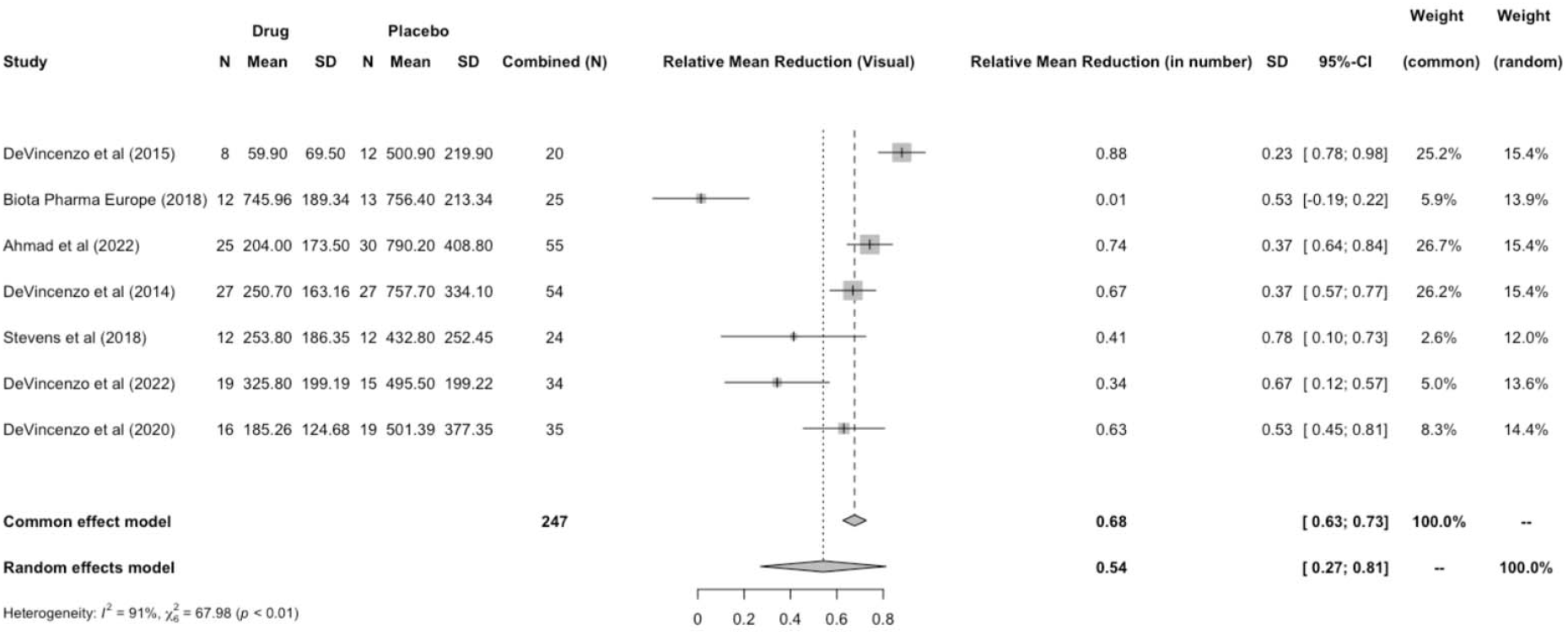
Forest plot for relative mean reduction (RMR) of mean viral load area under the curve (VL AUC). Note: VL is assessed using RT-qPCR assay. Studies report the unit of VL in log10 copies/mL or log10 PFUe/mL, where log10 PFUe/mL = log (base 10) plaque-forming unit equivalents per millilitre. In our analysis, we assumed a conversion factor of 1 across the units which is confirmed by hVIVO. Unit of VL AUC is log10 PFUe.hr/mL, where the symbol (.) denotes multiplication.

#### Relative mean reduction (RMR) of total symptom score area under the curve (TSS AUC)

This analysis involved 4 studies, ranging in size from 20 to 55 participants, a total of 144 participants were included. Individually, the RMR of TSS AUC in the treatment group compared to placebo varied from 35% to 82% (Figure 3). The I^2^ statistic was used to assess the heterogeneity across studies and was equal to 21% (p = 0.28, Chi-squared test with 3 degrees of freedom), indicating low heterogeneity across the studies (Figure 3). As a result, both the fixed effect and random effects models produced the same estimates of the RMR of mean TSS AUC in the treatment group compared to placebo, which was 76% (95% CI: 61% - 91%) (Figure 3).

**Figure 3.**
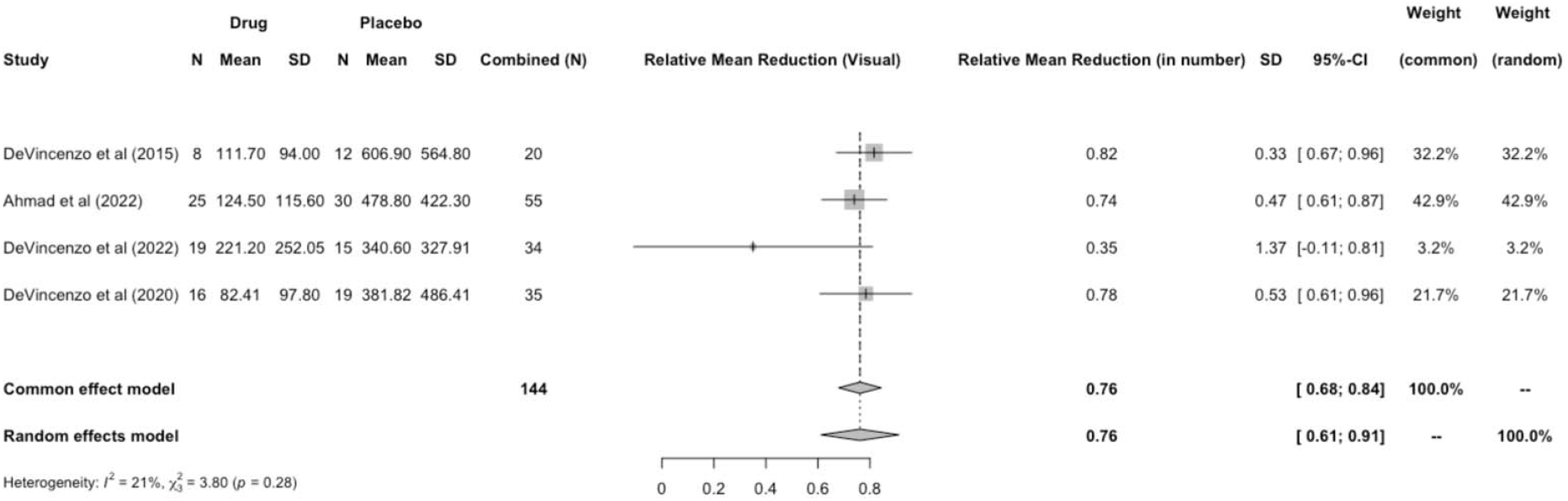
Forest plot for relative mean reduction (RMR) of mean total symptom score area under the curve (AUC). Note: The total symptom score is evaluated based on 10 clinical symptoms from the symptom dairy card. The reported AUC from each study is based on a window length of approximately 12-13 days.

### Secondary outcomes

#### Mean placebo VL AUC

This analysis included 7 studies ranging in size from 12 to 30 participants, with a total of 128 participants. Individually, the placebo mean VL AUC varied from 432.80 to 790.20 log_10_PFUe.hr/mL (Supplementary Figure 2). The I^2^ statistic was used to assess the heterogeneity across studies and was equal to 82% (p < 0.01, Chi-squared test with 6 degrees of freedom), indicating a considerable degree of heterogeneity (Supplementary Figure 2). Under a random effects model, the placebo mean VL AUC was 605.98 log_10_PFUe.hr/mL (95% CI: 491.60 – 720.37 log_10_PFUe.hr/mL) (Supplementary Figure 2).

#### Mean placebo VL at peak

This analysis included 7 studies ranging in size from 12 to 30 participants, with a total of 128 participants. Individually, the mean placebo VL at peak varied from 4.7 to 6.24 log_10_PFUe/mL (Supplementary Figure 3). The I^2^ statistic was used to assess the heterogeneity across studies and was equal to 55% (p = 0.04, Chi-squared test with 6 degrees of freedom), indicating a moderate degree of heterogeneity (Supplementary Figure 3). Under a random effects model, the mean placebo VL at peak was 5.38 log_10_PFUe/mL (95% CI: 4.94 – 5.81 log_10_PFUe/mL) (Supplementary Figure 3).

#### Time (in days) to mean placebo peak VL

Since most of the studies reported time to mean peak VL rather than mean time to peak VL for placebo subjects, we summarised the descriptive statistics for time to mean peak VL, which are evaluated assuming each study serves as a random subject. This analysis involved 6 studies, ranging in size from 12 to 30 participants, a total of 124 participants were included. Individually, the time to mean placebo peak VL from inoculation ranged from 6.2 to 7.75 days. The mean and median times to mean placebo peak VL were 6.99 days (95% CI: 6.24 – 7.74 days) and 6.74 days (95% CI: 6.2 – 8 days), respectively. In comparison to other viral diseases, time to mean peak VL occurs much earlier for influenza A/Wisconsin/67/2007 (H3N2) (2 days)^13^ and at a similar time in the disease course for SARS-CoV-2 B/Victoria/01/2020 (∼7.3 days)^14^. Similar time to mean peak VL (∼2 days) for influenza was reported in Carrat F, et.al.^35^ However, the time was evaluated using summary curves of viral shedding averaged over influenza types and subtypes, namely influenza A/Texas/36/91(H1N1), influenza A/Kawasaki/86(H1N1), influenza A/California(H1N1); influenza A/Victoria/3/75(H3N2), influenza A/Bethesda/1/85 (H3N2), influenza A/England/42/72(H3N2); and influenza B/Yamagata/16/88.

#### Mean placebo total symptom score area under the curve (TSS AUC)

This analysis involved 4 studies, ranging from 12 to 30 participants, 76 participants in total were included. Individually, the mean placebo TSS AUC varied from 340.60 to 606.90 (Supplementary Figure 4). The I^2^ statistic was used to assess the heterogeneity across studies and was equal to 0% (p = 0.41, Chi-squared test with 3 degrees of freedom), indicating no heterogeneity (Supplementary Figure 4). Under a random-effects model, the placebo mean TSS AUC was 426.53 (95% CI: 275.55 - 577.51) (Supplementary Figure 4).

#### Time (in days) to mean placebo peak total symptom score (TSS)

We report the descriptive statistics, as most of the studies reported time to mean peak TSS instead of mean time to peak TSS for placebo subjects. The statistics were evaluated analogously to those described above for VL. This analysis involved 5 studies, ranging from 12 to 30 participants, 107 participants in total were included. Individually, the time to mean placebo peak TSS from inoculation varied from 6 to 8.04 days. The mean and median times to mean placebo peak TSS were 7.09 days (95% CI: 6.06 – 8.12 days) and 6.87 days (95% CI: 6 – 8.04 days), respectively. The results show that the time to mean peak VL and mean peak symptom score are similar in RSV across multiple challenge studies, as has been observed in a non-interventional individual study^34^.

In comparison to other viral diseases, peak TSS occurs earlier for influenza A/Wisconsin/67/2007/H3N2 (3 days)^13^ and later in the disease course for SARS-CoV-2 B/Victoria/01/2020 (∼9.4 days)^14^. A similar time to mean peak TSS (∼3 days) for influenza was reported in Carrat F, et.al.^35^ However, the time was evaluated using summary curves of total symptom scores averaged over influenza types and subtypes, namely influenza A/Texas/36/91(H1N1), influenza A/Kawasaki/86(H1N1), influenza A/California(H1N1); influenza A/Victoria/3/75(H3N2), influenza A/Bethesda/1/85 (H3N2), influenza A/England/42/72(H3N2); and influenza B/Yamagata/16/88.

### Prediction Interval for a future HIC and Reference Interval for an individual recruited in a future HIC

We report the prediction interval and reference range for placebo VL AUC, placebo VL at peak and placebo TSS AUC. From Supplementary Table 2, the 95% prediction interval for mean placebo VL AUC, mean placebo VL at peak and mean placebo TSS AUC in a future HIC are (218.97 – 993.0 log_10_PFUe.hr/mL), (4.13 – 6.63 log_10_PFUe/mL) and (0 – 892.11), respectively. From Supplementary Table 3, based on the empirical approach, the 95% reference interval or the “normal” range where 95% of the measurements from individuals recruited in a future HIC would lie are (0 – 1335.30 log_10_PFUe.hr/mL), (1.55 – 9.24 log_10_PFUe/mL) and (0 – 1316.75 score.hr) for placebo VL AUC, placebo VL at peak and placebo TSS AUC, respectively. We have also provided the plots, including prediction intervals at the study-level and reference intervals at an individual-level (Supplementary Figure 5, Supplementary Figure 6, Supplementary Figure 7).

## Discussion

We conducted a systematic review and meta-analysis of RSV outcomes from human infection challenge trials, estimating effects in the placebo group and treatment effects on viral load (VL) and total symptom score (TSS) end-points. Where meta-analysis was not possible, we used descriptive statistics to estimate key time measures like time to peak VL and TSS. We included prediction intervals at the study-level and reference intervals at an individual-level for some of the measures. Our findings show RSV has a mean peak VL time similar to SARS-CoV-2, but later than influenza. Analysis of our primary outcomes demonstrated an average 54% reduction in viral load (VL) AUC, though with considerable variability across studies. Conversely, the total symptom score (TSS) AUC showed a larger average reduction of 76% and much lower heterogeneity, suggesting more consistent and reliable effects across studies for this outcome. For symptom dynamics, peak TSS occurs later in SARS-CoV-2 and earlier in influenza. In RSV, peak VL and TSS occur simultaneously, while in SARS-CoV-2 and influenza, peak TSS is delayed by 2 and 1 days, respectively.

RSV challenge studies typically assess RSV positivity from day 2 to day 5 post-inoculation before randomising participants to treatment. By day 5, all participants are assumed to be RSV positive and are randomised. However, a small proportion may test positive after randomisation, which may not fully reflect the clinical treatment scenario. The screening strategy used in the studies was designed to achieve an optimal infection rate. Despite this, some individuals do not become infected (defined as lab confirmed RSV infection) after inoculation. This rate varied between studies, thereby introducing possible heterogeneity in them. Some individuals may have been exposed to RSV in the intervening time between screening and inoculation and/or innate or cell-based immunity, which are not evaluated at screening, may be a better predictor of susceptibility than neutralising antibody levels alone. The heterogeneity in viral load in individuals that do become infected post inoculation may similarly be driven by the variable levels of RSV-specific as well as innate immunity in the healthy volunteer population. Imbalance in the viral load and symptom severity, prior infection timing across the underlying populations from challenge studies, seasonal differences across the studies and varying mechanisms of action of drugs across the challenge studies could be potential sources of heterogeneity, in addition to residual confounding due to unmeasured confounders.

There are several limitations in our study. The small number of studies may limit the statistical power of the results of the meta-analysis. Available RSV HIC data evaluate treatment response in healthy adults with known baseline RSV level seropositivity by controlled challenge strain that causes upper respiratory tract infection, limiting the evaluation of treatment response for lower respiratory tract infection. Findings from challenge trials may not be applicable to the general population, as participants are mainly young, healthy, White/Caucasian adults from the UK, whose responses may differ from those of other demographics or individuals with comorbidities. One of the key issues with many challenge studies is that they administer an antiviral experimental drug prior to the onset of symptoms. While this approach may show promise in controlled environments, it has not translated into clear clinical efficacy in real-world settings, since none of these experimental drugs tested in the challenge studies have been approved for RSV treatment. In actual clinical practice, patients typically begin treatment only after symptoms have appeared, which is often much later than in challenge studies. Limited data exist on viral load and symptom scores in vulnerable populations due to inconsistent sampling, especially before peak viral load and symptoms, and the lack of a validated symptom score tool. Therefore, results may be specific to the challenge strain (RSV-A Memphis-37b) and population studied. Viral load may have higher peaks and more protracted viremia in the young paediatric population. Future research should include under-represented ethnic/racial groups and individuals with comorbidities and investigate other strains like RSV-B. Other limitations include inconsistent outcome reporting, with only four of the seven studies reporting mean TSS AUC. Studies mostly reported time to mean peak VL/TSS, not mean time to peak, limiting our ability to perform a traditional meta-analysis.

Despite these limitations, our comprehensive assessment of the RSV HICs provides a detailed understanding of the VL and symptom score kinetics that will enhance understanding of RSV disease pathogenesis. Meta-analysis and descriptive statistics combined data across studies to assess placebo group characteristics and relative reduction, thereby increasing power and aiding investigators and sponsors in designing better challenge and symptomatic patient studies. These analyses provide insight into the heterogeneity in RSV viral load in healthy adults that may help to inform modelling and sample size estimations for studies in patient populations (at-risk adults and paediatrics) where such intense longitudinal viral load data are not available. The analysis using data from the placebo group can also inform the design of future challenge studies, enabling the use of fewer volunteers for the placebo group and incorporating historical external placebo data, thus making it cost-efficient. Reference intervals for some of the key measures will guide the clinician to determine if the patient’s measurement lies within the normal range, thus validating the patient’s inclusion in the HIC. Our study mainly focused on HICs that used RSV-A Memphis-37b as the challenge virus in the experimental stage, and therefore, the results from our study are applicable to RSV-A, which is the most prevalent strain. These results can be used to contextualise effects to other potential experimental drugs when tested on the same strain. Overall, findings from our analysis can help researchers and clinicians in significantly accelerating the development of RSV treatment interventions.

## Supporting information

Supplementar Online Content

## Data Availability

All data produced in the present work are contained in the manuscript.

## Contributors

Prosenjit Kundu: Literature search, data curation, formal analysis, methodology, software, supervision, validation, visualisation, data interpretation, writing-original draft.

Mark Quinn: Conceptualisation, literature search, data curation, formal analysis, validation, visualisation, writing-original draft.

Jared C. Christensen: Writing-review & editing.

Elaine Thomas: Data interpretation, writing-review & editing.

Sima S. Toussi: Literature search, data interpretation, writing-review & editing. Negar Niki Alami: Writing-review & editing.

Anindita Banerjee: Conceptualisation, literature search, data curation, supervision, data interpretation, writing-review & editing.

Prosenjit Kundu, Mark Quinn and Anindita Banerjee have accessed and verified the underlying data, including the original articles of both included and excluded studies, as well as the data extracted for the meta-analysis. All authors have read and approved the final version of the manuscript.

## Data Sharing Statement

This review is based on published data either from peer-reviewed manuscripts or from clinical trials register, and the data is publicly available. All the data and the code for meta-analysis can be accessed through GitHub link**: https://github.com/kundupro/RSV_HIC_meta_analysis**

## Acknowledgement

We thank Dr. Haitao Chu, Senior Director and Statistics Lead at Pfizer Inc., for providing insightful comments on the paper. We thank Pfizer Inc. for its support.

