## Supplementar Online Content for "Role of human infection challenge studies (HICs) in drug development for respiratory syncytial virus (RSV): Systematic Review and Meta-Analysis"

for

**Supplementary Table 1.** Infection rates reported in the studies for different subgroups.

**Supplementary Table 2.** Prediction interval for a future Human Challenge Trial (HCT).

**Supplementary Table 3.** Reference Interval for an individual recruited in a future HCT.

**Supplementary Figure 1.** Assessment of risk of bias for the selected HCTs.

**Supplementary Figure 2.** Forest plot for mean viral load area under the curve (VL AUC) in placebo.

**Supplementary Figure 3***.* Forest plot for mean viral load at peak in placebo.

**Supplementary Figure 4.** Forest plot for mean total symptom score area under the curve (TSS AUC) in placebo.

**Supplementary Figure 5.** Prediction Interval Plot for Mean Placebo VL AUC and Reference Interval Plot for Placebo VL AUC.

**Supplementary Figure 6.** Prediction Interval Plot for Mean Placebo VL at peak and Reference Interval Plot for Mean Placebo VL at peak.

**Supplementary Figure 7.** Prediction Interval Plot for Mean Placebo TSS AUC and Reference Interval Plot for Mean Placebo TSS AUC.

**Supplementary References.**

**Supplementary Table 1.** Infection rates reported in the studies for different subgroups.

| **Study** | **Drug** | **Infection^a^ rate** | | | |
| --- | --- | --- | --- | --- | --- |
|  |  | **Treatment + Placebo** | | **Treatment group** | |
|  |  | **Overall^b^** | **Prior treatment assignment^c^** | **Overall^b^** | **Prior treatment assignment^c^** |
| Ahmad et al (2022)^1^ | EDP-938 | 72.37% | Not reported | 65.79% | Not reported |
| Biota Pharma Europe (2018)^2^ | BTA-C585 | Not reported | Not reported | Not reported | Not reported |
| DeVincenzo et al (2014)^3^ | GS-5806 | Not reported | 69.23% | Not reported | 69.23% |
| DeVincenzo et al (2015)^4^ | ALS-008176 | 62.50% | Not reported | 57.14% | Not reported |
| DeVincenzo et al (2022)^5^ | PC786 | 73.21% | Not reported | 78.51% | Not reported |
| DeVincenzo et al (2020)^6^ | RV521 | 79.55% | 68.18% | 72.72% | 63.64% |
| Stevens et al (2018)^7^ | JNJ-53718678 | 70.56% | Not reported | 66.67% | Not reported |

**^a^** *Infection is defined as positive for lab confirmed RSV infection using RT-qPCR test.*

**^b^** Overall infection rate includes infections *evaluated prior to dosing(baseline) and at least at two occasions after baseline.*

*^c^* *Includes infections prior to dosing.*

**Supplementary Table 2.** Prediction interval for a future Human Challenge Trial (HCT).

| **Outcome** | **95% Prediction Interval^*^** |
| --- | --- |
| Mean placebo VL AUC  (log_10_PFUe.hr/mL) | (218.97 – 993.00) |
| Mean placebo VL at peak  (log_10_PFUe/mL) | (4.13 – 6.63) |
| Mean placebo TSS AUC (score.hr) | (217.98 – 635.08) |

*^*^Prediction interval is calculated using the formula:*$\hat{\mu}\pm t_{k-2}^{0.05}\sqrt{\left\{ \hat{\tau}^{2}+ \hat{SE\left( \hat{\mu} \right)^{2}} \right\}}$*, where* $\hat{\mu}$*is the meta-analysis estimate,* $\hat{SE(\hat{\mu})}$ *is the standard error of the meta-analysis estimate,* $\hat{\tau}^{2}$*is the between study variance,* $t_{k-2}^{0.05}$ *is the 97.5^th^ percentile of the t-distribution with k-2 degrees of freedom, k denoting the number of studies^8^.*

**Supplementary Table 3.** Reference Interval for an individual in placebo recruited in a future HCT.

| **Outcome** | **95% Reference Interval** | | |
| --- | --- | --- | --- |
|  | **Frequentist** | **Bayesian** | **Empirical** |
| VL AUC (log_10_PFUe.hr/mL) | (0 – 1295.54) | (0 – 1375.05) | (0 – 1335.30) |
| VL at peak (log_10_PFUe/mL) | (1.58 – 9.18) | (1.38 – 9.36) | (1.55 – 9.24) |
| TSS AUC (score.hr) | (0 – 1288.25) | (0 – 1519.80) | (0 – 1316.75) |

*Note: The lower bounds with negative values are truncated to 0 as the outcomes cannot take negative values. The symbol (.) in the unit of VL AUC (log_10_PFUe.hr/mL) and TSS AUC (score.hr) denotes multiplication.*

**Supplementary Figure 1.** Assessment of risk of bias for the selected HCTs.

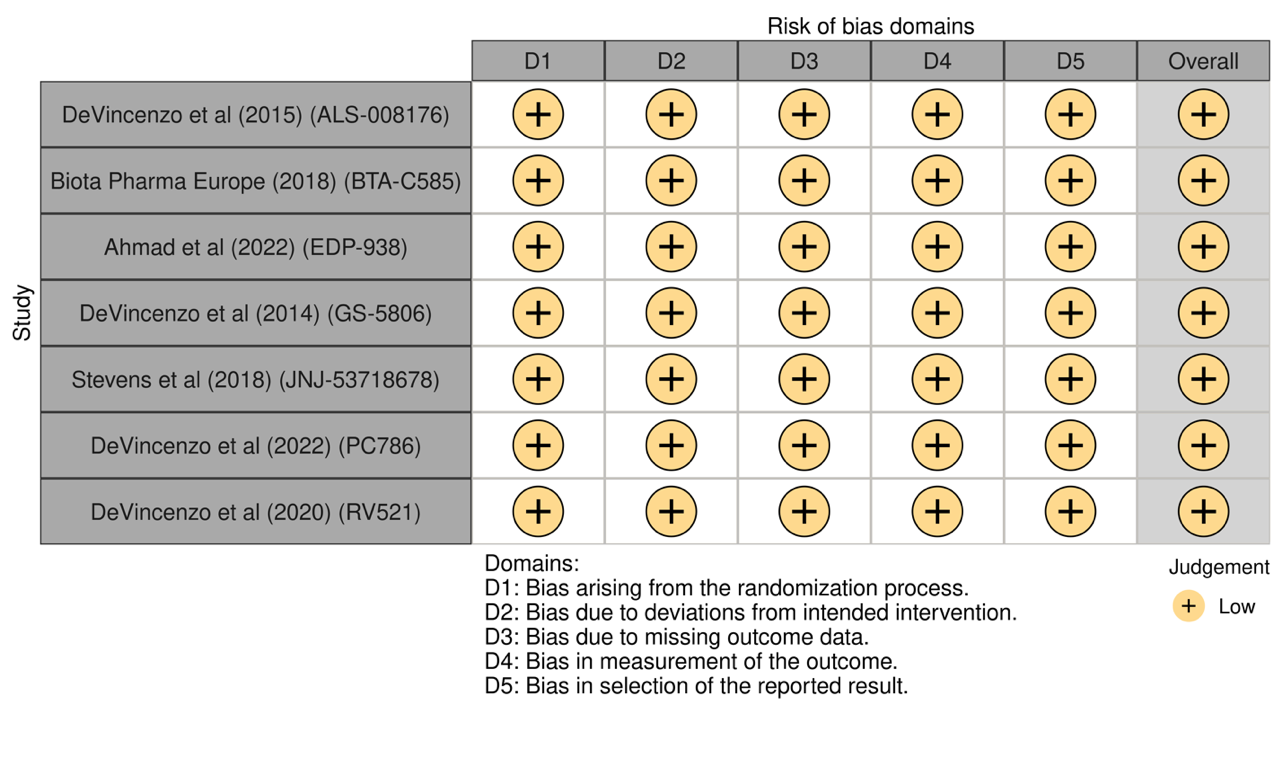

**Supplementary Figure 2.** Forest plot for mean viral load area under the curve (VL AUC) in placebo.
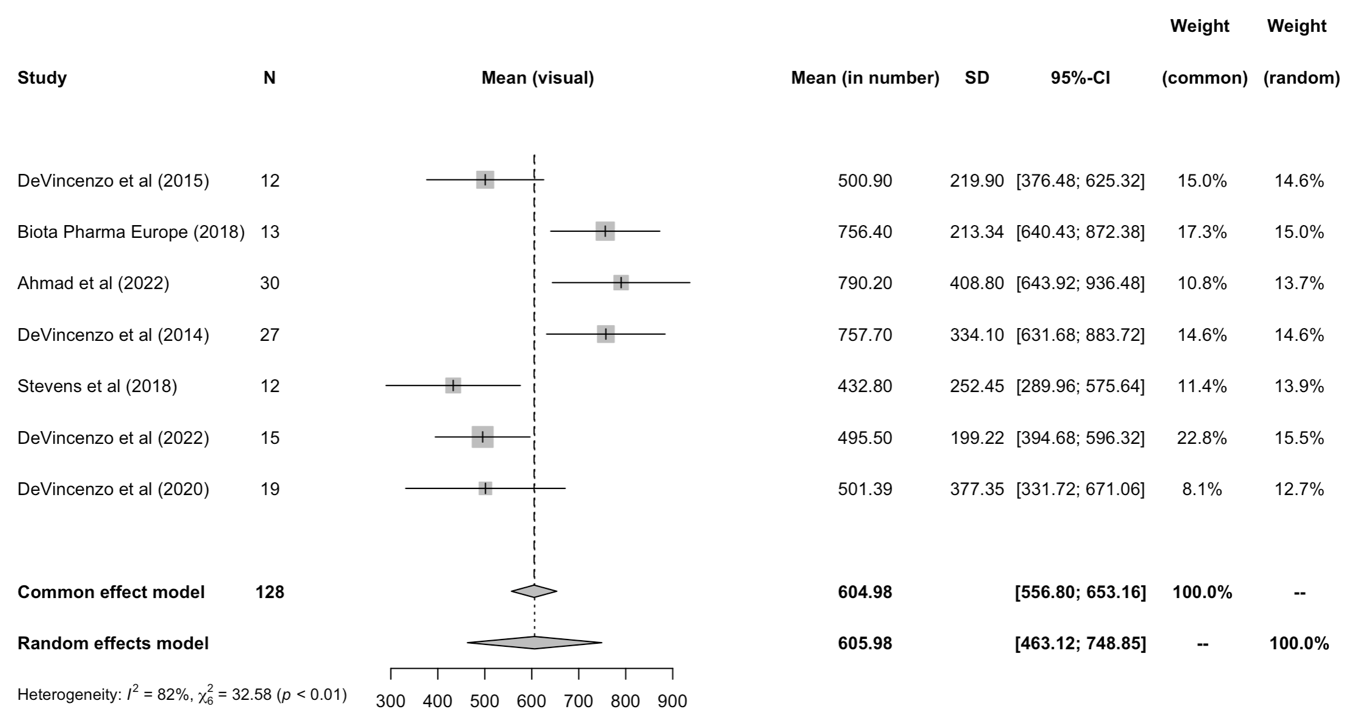

*Note: VL is assessed using RT-qPCR assay. Studies report the unit of VL in log10 copies/mL or log10 PFUe/mL, where log10 PFUe/mL = log (base 10) plaque-forming unit equivalents per milliliter. In our analysis, we assumed a conversion factor of 1 across the units and is confirmed from hVIVO. Unit of VL AUC is log10 PFUe.hr/mL, where the symbol (.) denotes multiplication.*

**Supplementary Figure 3.** Forest plot for mean viral load at peak in placebo**.**

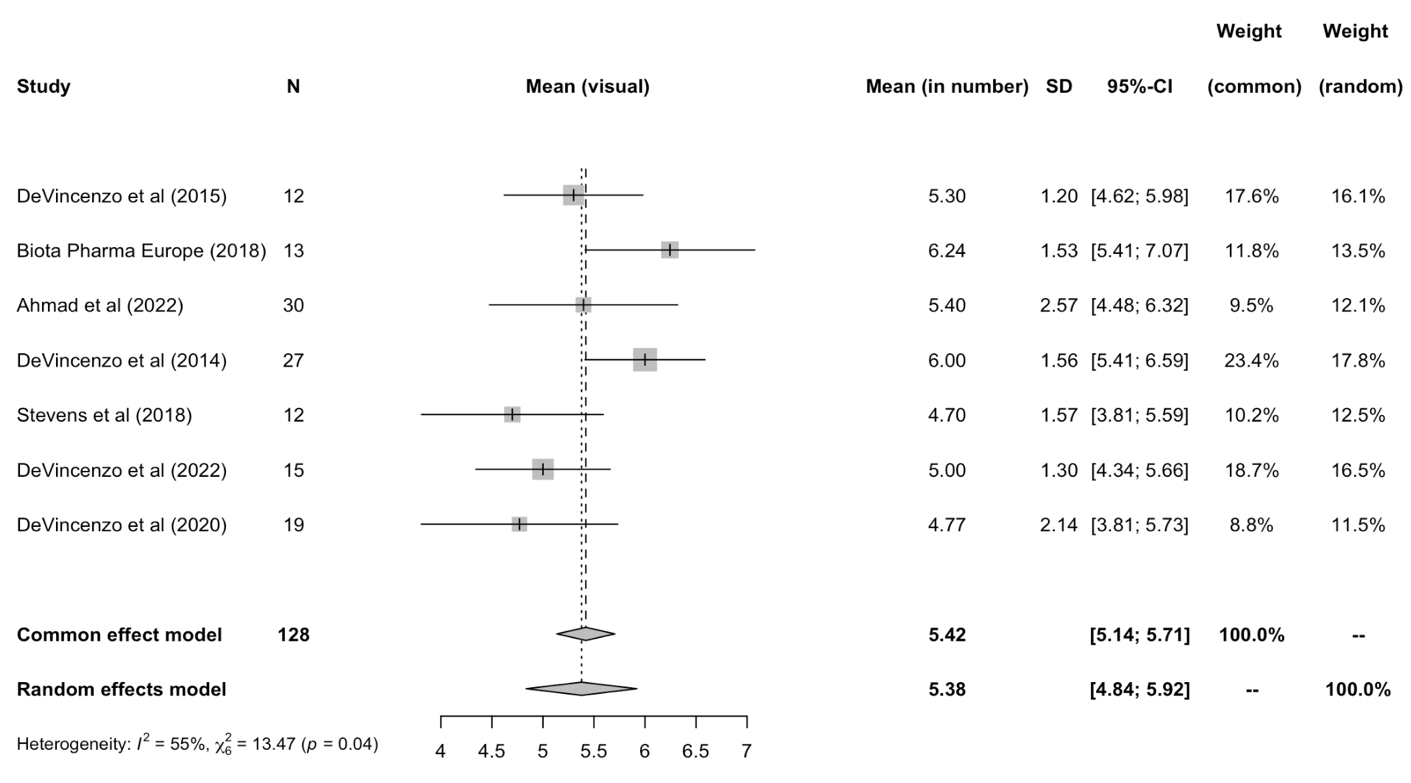

*Note: VL is assessed using RT-qPCR assay. Studies report the unit of VL in log10 copies/mL or log10 PFUe/mL, where log10 PFUe/mL = log (base 10) plaque-forming unit equivalents per milliliter. In our analysis, we assumed a conversion factor of 1 across the units and is confirmed from hVIVO.*

**Supplementary Figure 4.** Forest plot for mean total symptom score area under the curve (TSS AUC) in placebo.

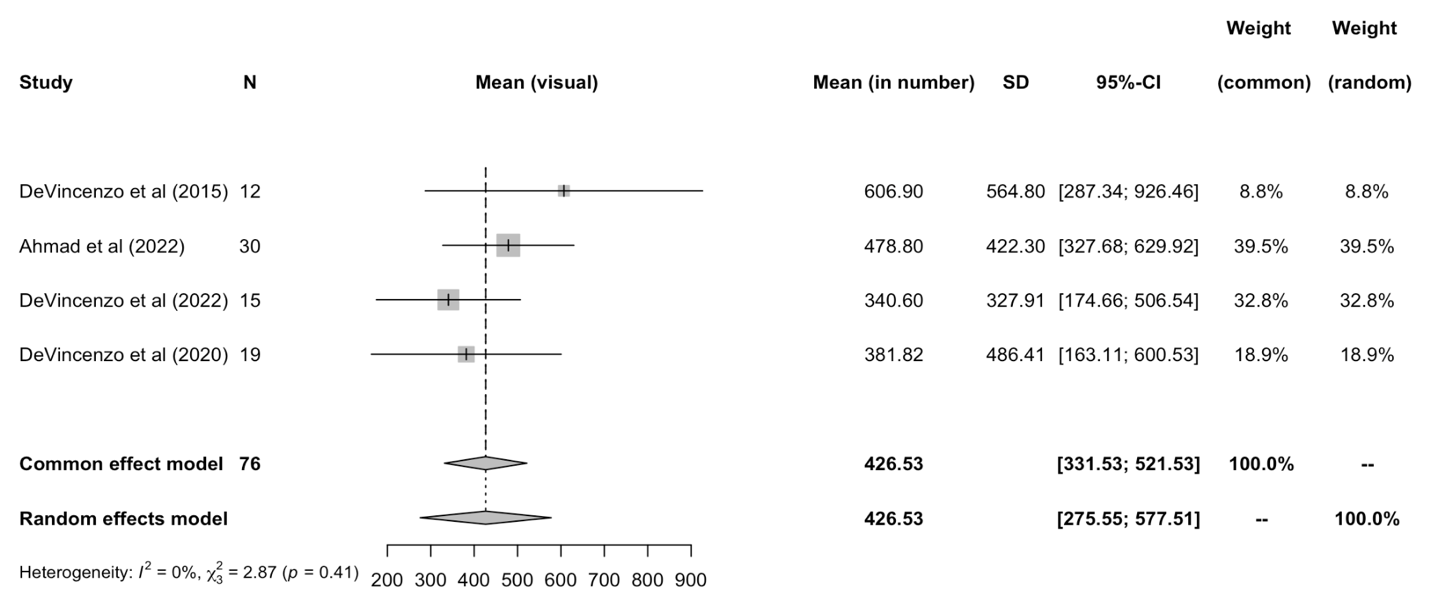

*Note: The total symptom score is evaluated based on 10 clinical symptoms from symptom dairy card. The reported AUC from each study is based on a window length of approximately 12-13 days.*

**Supplementary Figure 5.** Prediction Interval Plot for Mean Placebo VL AUC and Reference Interval Plot for Placebo VL AUC.

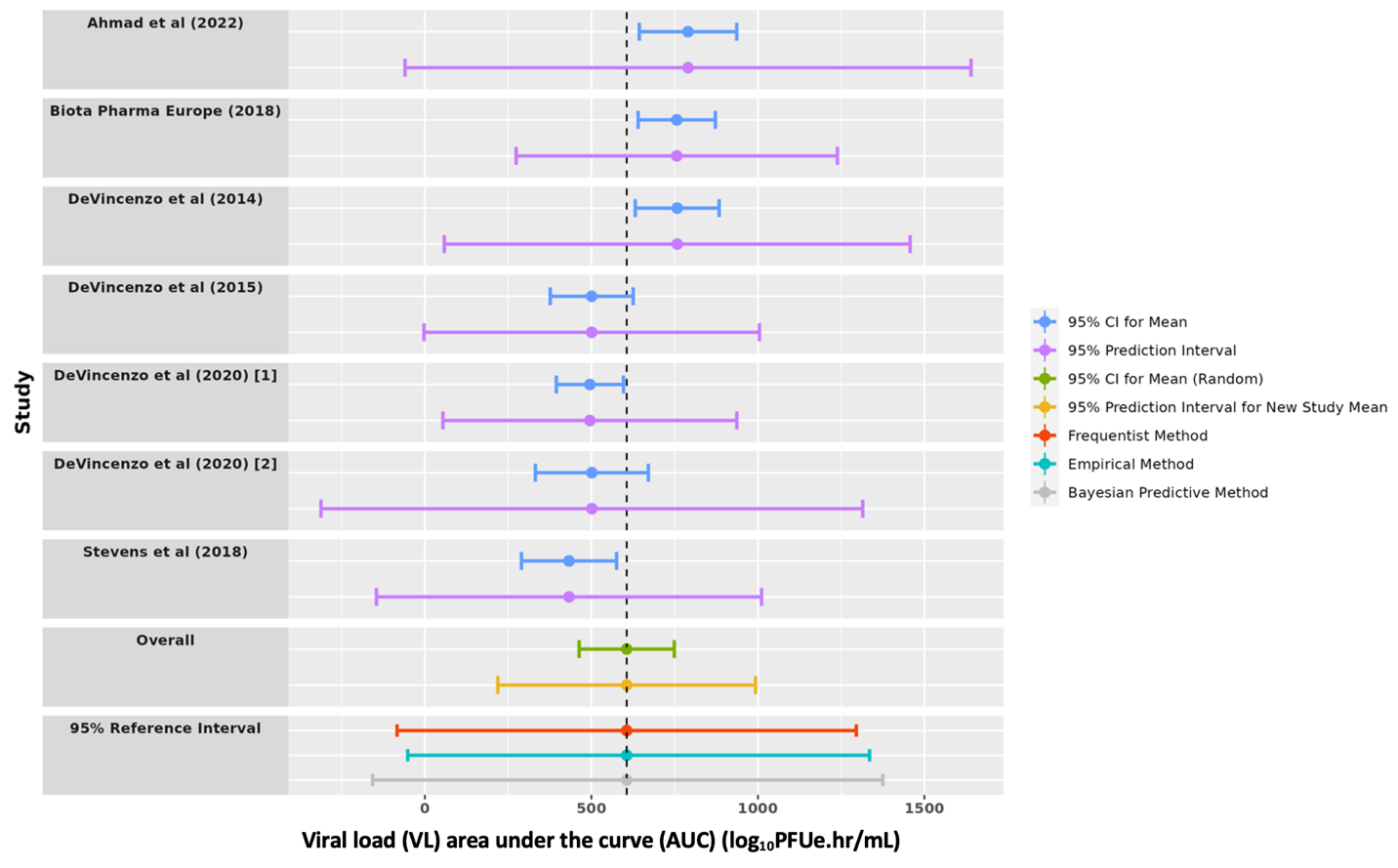

**Supplementary Figure 6.** Prediction Interval Plot for Mean Placebo VL at peak and Reference Interval Plot for Mean Placebo VL at peak.

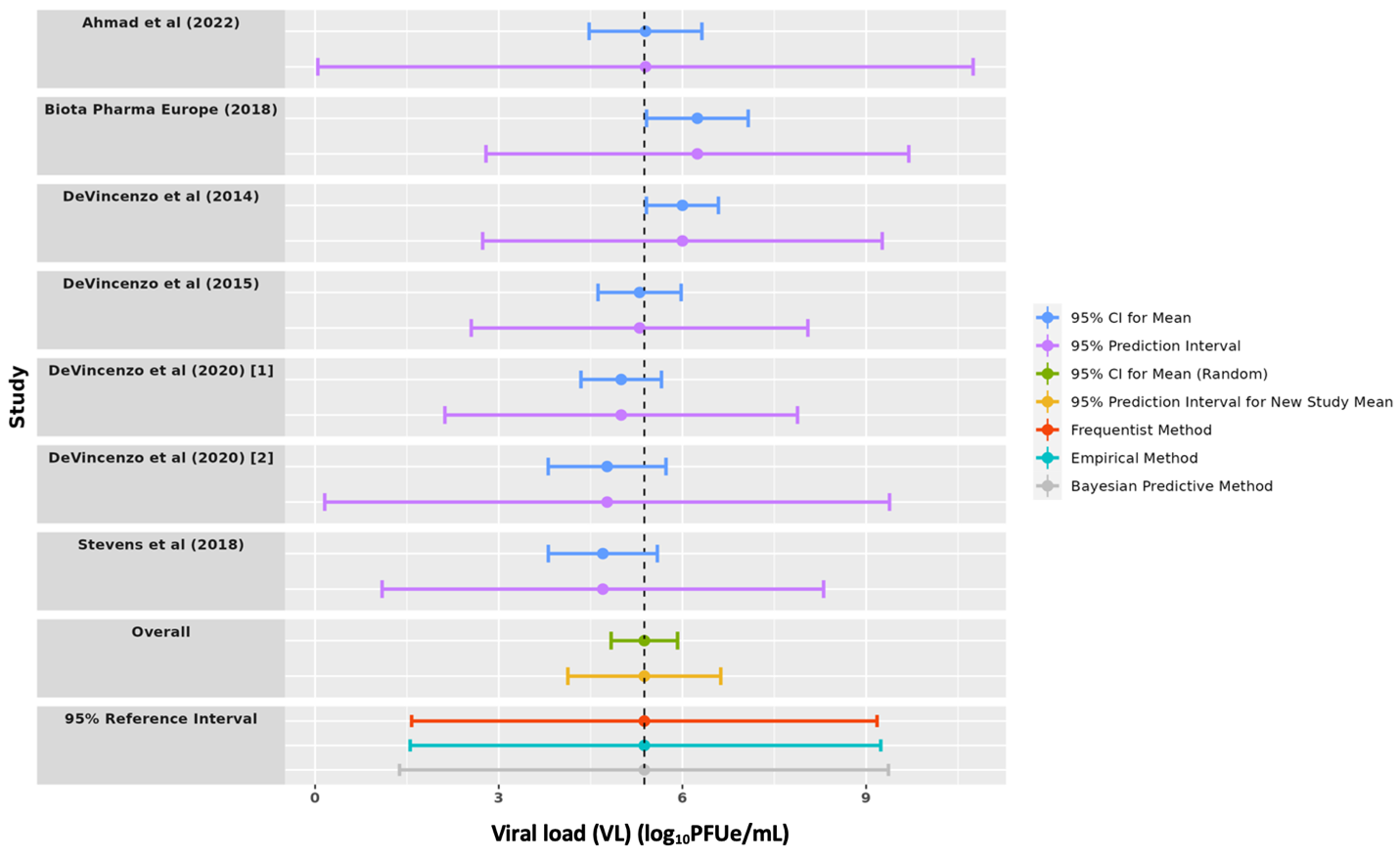

**Supplementary Figure 7.** Prediction Interval Plot for Mean Placebo TSS AUC and Reference Interval Plot for Mean Placebo TSS AUC.

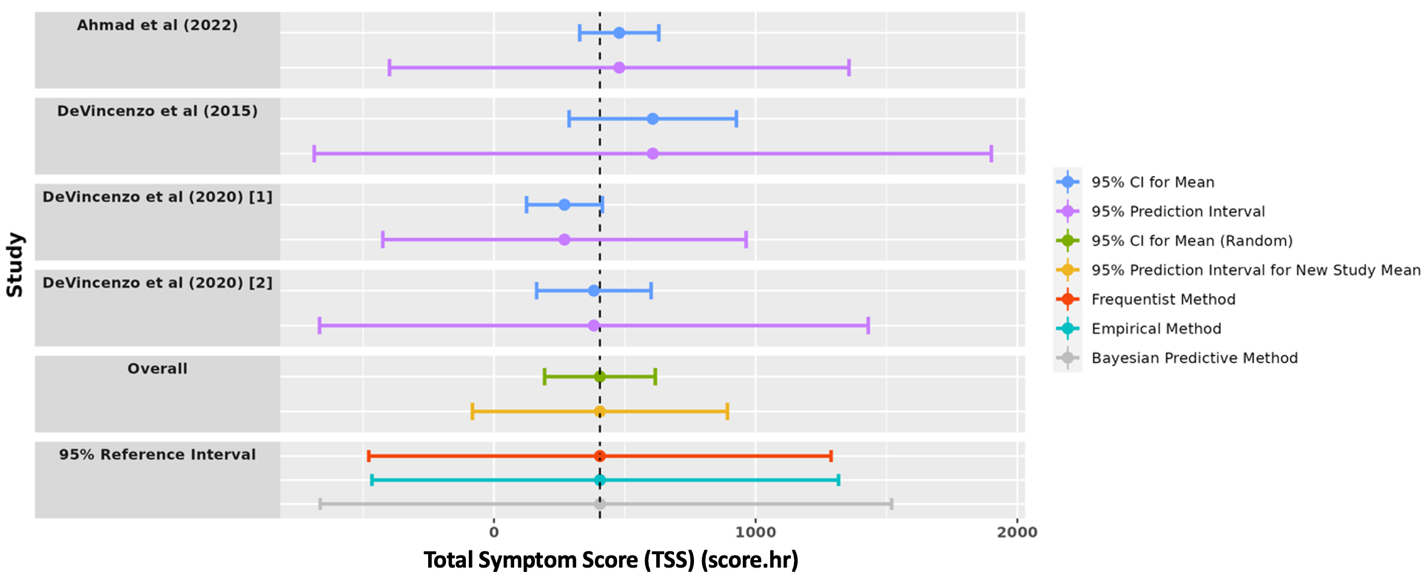
